# Comparison of infected and vaccinated transplant recipients highlights the role of Tfh and neutralizing IgG in COVID-19 protection

**DOI:** 10.1101/2021.07.22.21260852

**Authors:** Xavier Charmetant, Maxime Espi, Ilies Benotmane, Francoise Heibel, Fanny Buron, Gabriela Gautier-Vargas, Marion Delafosse, Peggy Perrin, Alice Koenig, Noelle Cognard, Charlene Levi, Floriane Gallais, Louis Manière, Paola Rossolillo, Eric Soulier, Florian Pierre, Anne Ovize, Emmanuel Morelon, Thierry Defrance, Samira Fafi-Kremer, Sophie Caillard, Olivier Thaunat

**Affiliations:** CIRI, INSERM U1111, Université Claude Bernard Lyon I, CNRS UMR5308, Ecole Normale Supérieure de Lyon, Univ. Lyon, 21 avenue Tony Garnier, 69007 Lyon, France; Department of Nephrology and Transplantation, Strasbourg University Hospital, Strasbourg, France; Department of Virology, Strasbourg University Hospital, Strasbourg, France; Inserm UMR S1109, LabEx Transplantex, Fédération de Médecine Translationnelle de Strasbourg (FMTS), Université de Strasbourg, Strasbourg, France; Hospices Civils de Lyon, Edouard Herriot Hospital, Department of Transplantation, Nephrology and Clinical Immunology, 5, place d’Arsonval, 69003 Lyon, France; Claude Bernard University (Lyon 1), 43 boulevard du 11 Novembre 1918, 69622 Villeurbanne France; Institut de Génétique et de Biologie Moléculaire et Cellulaire (IGBMC), Centre National de la Recherche Scientifique (CNRS), UMR 7104, Institut National de la Santé et de la Recherche Médicale (INSERM), U1258, Université de Strasbourg, Illkirch, France; Eurofins Biomnis Laboratory, 69007 Lyon, France

## Abstract

Transplant recipients, which receive therapeutic immunosuppression to prevent graft rejection, are characterized by high COVID-19-related mortality and defective response to vaccines. Having observed that previous infection by SARS-CoV-2 but not the standard “2 doses” scheme of vaccination, provided complete protection against COVID-19 to transplant recipients, we undertook this translational study to compare the cellular and humoral immune responses of these 2 groups of patients. Neutralizing anti-Receptor Binding Domain (RBD) IgG were identified as the critical immune effectors associated with protection. Generation of anti-RBD IgG was dependent upon spike-specific T follicular helper (Tfh) CD4+ T cells, which acted as limiting checkpoint. Tfh generation was impeded by high dose mycophenolate mofetil in non-responders to vaccine but not in infected patients, suggesting that increasing immunogenicity of vaccine could improve response rate to mRNA vaccine. This theory was validated in two independent prospective cohorts, in which administration of a 3^rd^ dose of vaccine resulted in the generation of anti-RBD IgG in half of non-responders to 2 doses.

**One sentence summary:** The generation of neutralizing IgG, which protects kidney transplant recipients from COVID-19, requires T follicular helper cells.

## Introduction

In December 2019, an outbreak of apparently viral pneumonia of unknown etiology emerged in the city of Wuhan, in the Chinese province of Hubei (*1*). On 9 January 2020, the World Health Organization (WHO) announced the discovery of a novel coronavirus officially named SARS-CoV-2, which is the pathogen responsible for this infectious respiratory disease called COVID-19 (CoronaVIrus Disease). The disease quickly disseminated from Wuhan and as at 8 July 2021, more than 184 million cases have been confirmed in 214 countries (*2*), leading the WHO to consider COVID-19 as the first pandemic triggered by a coronavirus.

Among the various alarms raised by the pandemic was its impact on the population of transplanted patients, whose COVID-19-related mortality was estimated ∼20%, several magnitudes higher than that of the general population (*3–7*). This vulnerable population of patients was therefore prioritized for vaccination against SARS-CoV-2 by health authorities (*8*). However, prevention of allograft rejection requires life-long immunosuppression regimens, which non-specifically inhibit T and B cells in transplant recipients, resulting in reduced response rates to vaccines (*9, 10*). As expected, several recent publications have documented that immunosuppressed transplant recipients develop mitigated immune responses following the “standard” 2 doses scheme of vaccination with any of the 2 approved SARS-CoV-2 mRNA vaccines (*11–14*).

Although insufficiency of vaccinal protection in transplant recipients has emerged as a concern due to accumulating reports of severe COVID-19 in vaccinated patients (*15, 16*), the underlying immune mechanisms explaining this problem are still elusive (*17*). In an attempt to determine the relative contribution of humoral and T cell immunity in conferring protection against COVID-19 and understand immunosuppression-induced defects following SARS-CoV-2 vaccination, we undertook a prospective translational study that compared infected and vaccinated transplant recipients.

## Results

### Infection but not vaccination confers full protection against COVID-19 to transplant recipients

The incidence of COVID-19 was monitored in all 889 renal transplant recipients of Strasbourg University Hospital and compared between those with previous history of infection with SARS-CoV-2 (group “infected”, n=153) and those who received the 2 doses “standard” scheme of vaccination with mRNA-1273 (group “vaccinated”, n=736; **Figure 1A**). Strikingly, while none of the infected patients did develop reinfection, 15 vaccinated patients (0% vs 2.04%, p=0.037) developed COVID-19 (**Figure 1B**). Of note this observation was made while the follow-up period of infected patients was significantly longer than that of vaccinated patients (243+/-161 vs 102+/-18 days, p<0.0001; **Figure 1A**).

**Figure 1:**
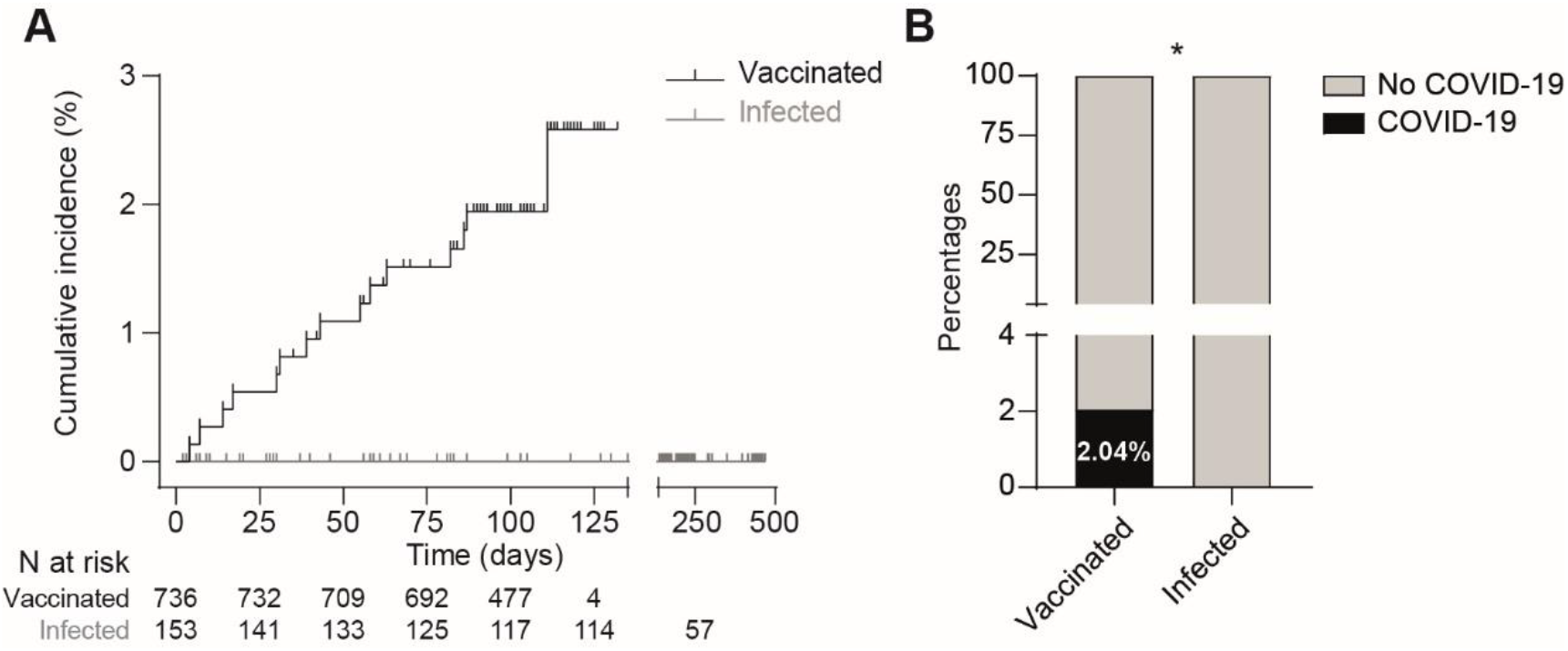
Infection but not vaccination confers full protection against COVID-19 to transplant recipients. Protection against COVID-19 was compared between renal transplant recipients with previous history of infection with SARS-CoV-2 (group “infected”, grey curve) and those who received the 2 doses “standard” scheme of vaccination with mRNA-1273 (group “vaccinated”, black curve). **A.** Cumulative incidence in the two groups was plotted using the Kaplan–Meier method. Log-rank test, p=0.06. **B.** Percentage of patients who developed COVID-19 in the 2 groups was compared. Chi-square test, *: p≤0.05.

We concluded that in contrast with vaccination, SARS-CoV-2 infection confers full protection against COVID-19 to immunocompromised transplant recipients.

### Population of the mechanistic study

Comparison of cellular and humoral immune responses developed by infected and vaccinated transplant patients offers a unique opportunity to determine which immune effector(s) are associated with protection against COVID-19 in this vulnerable population (*3–7*).

The COVATRHUS cohort was therefore established to prospectively collect synchronous serum and PBMCs samples from renal transplant recipients diagnosed with COVID-19 in absence of previous vaccine injection (group “infected”, n=21) or vaccinated with 2 doses of mRNA-1273 (group “vaccinated”, n=29). The clinical characteristics of the cohort are presented **Table 1**. Briefly, the 2 groups had similar profiles, except for lymphocytes and monocytes counts that were lower in infected patients (**Table 1**). Of note, the severity of COVID-19 in infected patients was mainly mild/moderate (16/21, 76%; **Table 2**) and most of them did not require hospitalization (14/21, 67%; **Table 2**).

**Table 1.** Characteristics of patients from COVATRHUS cohort.

| n (%) or median [IQR] | Vaccinated<br>N = 29 | Infected<br>N = 21 | p* |
| --- | --- | --- | --- |
| **Age (y)** | 56.4 [43.4; 68.8] | 55.0 [36.9; 60.5] | 0.359 |
| **Male** | 18 (62) | 13 (62) | 0.991 |
| **Time from transplantation (y)** | 6.96 [2.1; 16.1] | 2.0 [1.4; 7.5] | **0.049** |
| **Immunosuppressive drugs** |  |  |  |
| CNI | 28 (97) | 19 (90) | 0.565 |
| MMF/MPA | 22 (76) | 20 (95) | 0.117 |
| Steroids | 19 (66) | 14 (67) | 0.933 |
| imTOR | 3 (10) | 1 (5) | 0.473 |
| Belatacept | 1 (3) | 1 (5) | 0.815 |
| **Biological data** |  |  |  |
| Lymphocytes (G/L) | 1.24 [1.00; 1.87] | 1.03 [0.64; 1.33] | **0.024** |
| Monocytes (G/L) | 0.55 [0.41; 0.78] | 0.43 [0.35; 0.55] | **0.036** |
| CRP (mg/L) | 4.0 [4.0; 5.5] | 5.2 [4.0; 40.1] | 0.065 |
| Albumin (g/L) | 44 [43;46] | 43 [41; 47] | 0.563 |
| Creatinine (μmol/L) | 132 [101; 190] | 100 [78; 138] | 0.057 |

\*: qualitative variables were compared using a Chi-square test, quantitative variables were compared using a Mann-Whitney test.
**Abbreviations:** IQR: interquartile range; y, years; CNI: calcineurin inhibitor; MMF/MPA: mycophenolate mofetil/mycophenolic acid; imTOR, inhibitor of the mechanistic target of rapamycin.

**Table 2.** Characteristics of COVID-19 of infected patients.

| n (%) | Infected<br>N = 21 |
| --- | --- |
| **Disease severity** |  |
| Asymptomatic | 2 (10) |
| Mild | 12 (57) |
| Moderate | 4 (19) |
| Severe | 3 (14) |
| Critical | 0 (0) |
| **Management** |  |
| Hospitalization | 7 (33) |
| Outpatient | 14 (67) |
| **Oxygen requirement** | 4 (19) |
| **Drug therapy** |  |
| Antibiotics | 6 (29) |
| Remdesivir | 1 (5) |
| High dose steroids | 3 (14) |
| **Changes in immunosuppression** |  |
| Cessation of CNi | 2 (10) |
| Cessation of MMF/MPA | 7 (33) |

**Abbreviations:** CNi, calcineurin inhibitor; MMF/MPA: mycophenolate mofetil/mycophenolic acid.

### Similar cellular immunity in infected and vaccinated transplant recipients

Virus-specific CD8+ T cells reduce disease severity and promote recovery in many respiratory infections, including those driven by coronaviruses (*23, 24*), by eliminating infected cells (virus “factories”). Optimal generation of these cytotoxic effectors depends upon the help provided by the Th1 CD4+ T cells (*25*).

Cytotoxic CD8+ T cells directed against the spike protein of SARS-CoV-2, identified by the co-expression of CD69 and CD137 (*20*), could be detected in the circulation of both vaccinated and infected patients (**Figure 2A**). However, only the latter had CD8+ T cells directed against the other proteins of the virus (nucleocapsid and membrane). This finding was expected since nucleocapsid and membrane proteins are not included in the vaccine formulation (**Figure 2A & 2B**). Despite a trend for more spike-specific CD8+ T cells in the circulation of infected patients, the difference with the vaccinated group did not reach statistical significance (**Figure 2B**), even when all specificities (spike, nucleocapsid and membrane) were added together to better take into account the difference of repertoire size between the 2 groups (**Figure 2C**). The exact same observations were made when the analysis was focused on Th1 subset of CD4+ T cells (**Figure 2D-F**).

**Figure 2:**
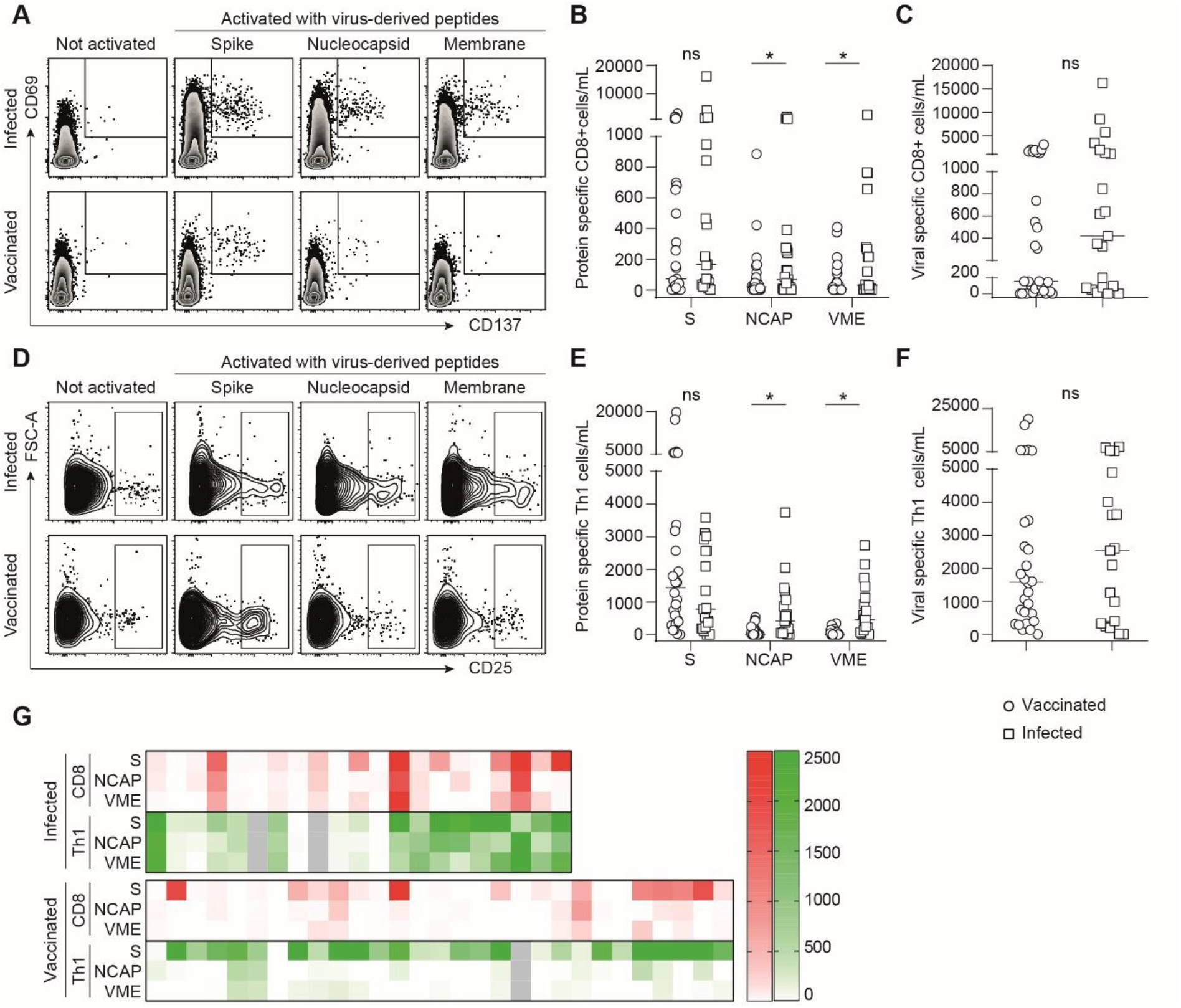
Anti-SARS-CoV-2 specific cellular immunity in infected and vaccinated transplant recipients. T cells directed against the various proteins [Spike (S), Nucleocapsid (NCAP), and membrane (VME)] of SARS-CoV-2 were enumerated in the circulation of infected (open square) and vaccinated (open circle) transplant recipients. The bar indicates the median. Mann-Whitney test; ns, p>0.05; *, p≤0.05. **A-C** Enumeration of SARS-CoV-2-specific CD8+ cytotoxic T cells. **A.** Flow cytometry profiles of a representative patient of each group are shown. **B.** The count of CD8+ cytotoxic T cells specific of each viral protein is plotted for each patient. **C.** For each patient, the total number of virus-specific CD8+ cytotoxic T cells is plotted. **D-F** Enumeration of SARS-CoV-2-specific Th1 CD4+ helper T cells. **D.** Flow cytometry profiles of a representative patient of each group are shown. **E.** The count of Th1+ CD4+ T cells specific of each viral protein is plotted for each patient. **F.** For each patient, the total number of virus-specific Th1+ CD4+ T cells is plotted. **G.** The global repertoire of anti-SARS-CoV-2 cellular response (CD8, red; Th1, green) is compared between infected (upper row) and vaccinated (lower row) transplant recipients.

We concluded that although the repertoire of the cellular immune response directed against SARS-CoV-2 is wider in infected patients (**Figure 2G**), the resulting small (and non-significant) increase in cellular effectors is unlikely to account alone for the drastic advantage in term of protection observed as compared with vaccinated transplant recipients (**Figure 1**). Another strong argument in favor of this hypothesis is the fact that some infected patients (**Figure 2G**) had no detectable virus-specific T cells, suggesting that their protection was due to other types of immune effectors.

### Presence of neutralizing IgG correlates with protection against COVID-19 in transplant recipients

Beside cellular effectors, the adaptive immune system also generates antibodies against SARS-CoV-2.

As expected, antibodies directed against viral nucleocapsid (not included in the vaccine formulation) were exclusively detected in patients from the infected group (**Figure 3A**), but only in half of them (11/21, 52%). In contrast almost all (20/21, 95%) infected transplant recipients developed anti-RBD IgG (**Figure 3B**). The spike glycoprotein mediates virus entry into target cells via the ACE2 receptor and it has been shown that antibodies directed against the RBD can block viral infection of human cells in vitro and counter viral replication in vivo (*26–30*). In line with these studies, and despite the fact that the titers were lower than those observed in a cohort of 30 vaccinated healthy volunteers (*31*), sera of infected transplant recipients still very efficiently block pseudo-virus entry in human cells *in vitro* (**Figure 3C**). A positive correlation between anti-RBD IgG titer and the result of the in vitro neutralization assay was demonstrated (**Figure 3D**).

**Figure 3:**
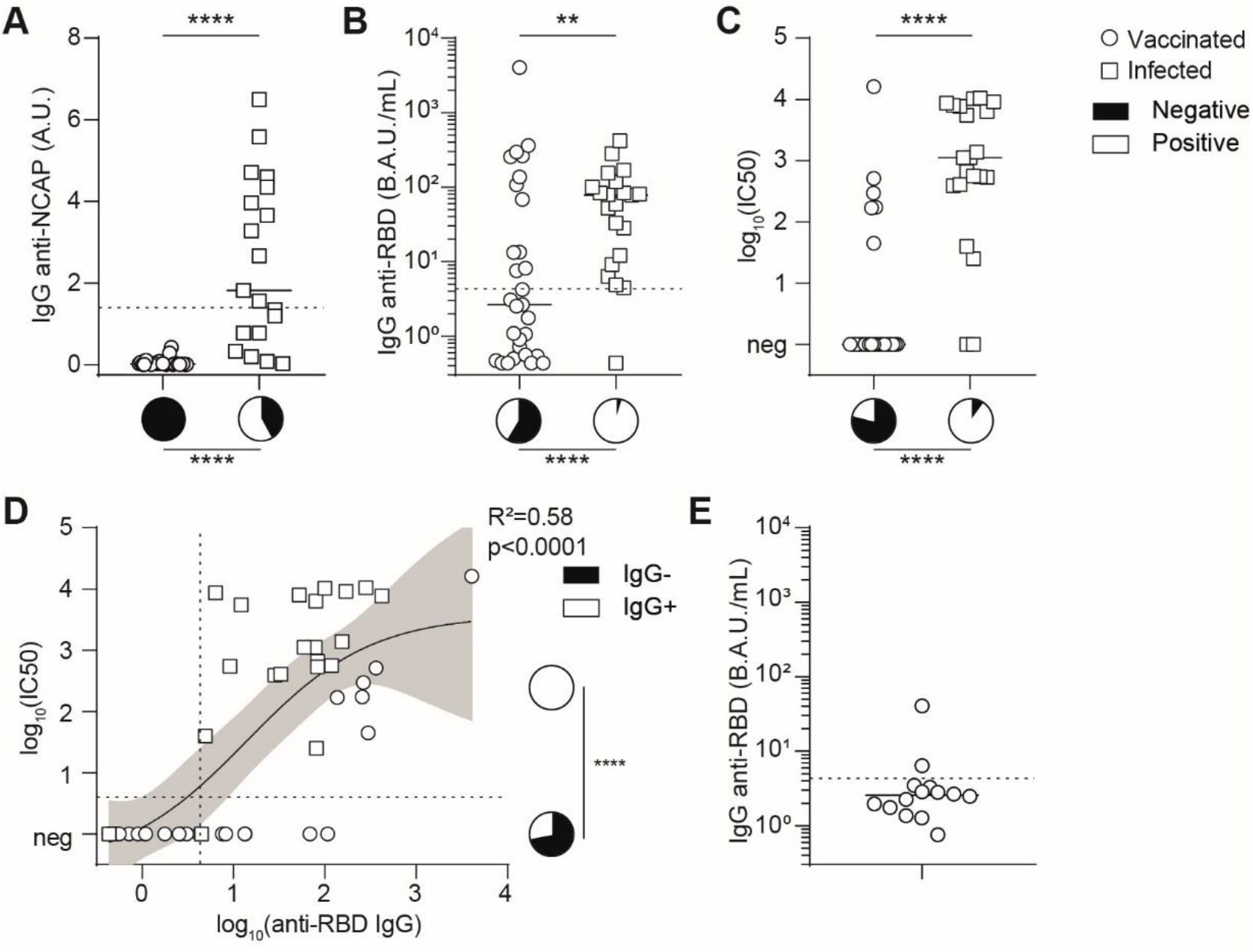
Anti-SARS-CoV-2 specific humoral immunity in infected and vaccinated transplant recipients. **A-B.** The titers of antibodies directed against the various proteins of SARS-CoV-2 were measured in the circulation of infected (open square) and vaccinated (open circle) transplant recipients. Dotted line indicates the threshold of positivity of the assay. The bar indicates the median. Pie charts are used to compare proportions. Mann-Whitney test for comparison of antibody titers and Chi-square for comparison of proportions; *, p≤0.05; ***,p<0.001; ****, p<0.0001. **A.** Titers of anti-Nucleocapsid (NCAP) IgG. **B.** Titers of anti-Receptor-Binding Domain of spike (RBD) IgG. **C.** The neutralizing capacity of patients’ serum was compared between infected (open square) and vaccinated (open circle) transplant recipients. The bar indicates the median. Pie charts are used to compare proportions. Mann-Whitney test for comparison of neutralizing titers and Chi-square for comparison of proportions; ****, p<0.0001. **D.** The logical relation between anti-RBD IgG titer and the neutralizing capacity of the serum is shown. The pie charts represent the proportion of patients without (black) or with (white) anti-RBD IgG. Chi-square test; ****, p<0.0001. **E**. The titers of anti-RBD IgG were measured in the circulation of patients who developed COVID-19 after vaccination. Dotted line indicates the threshold of positivity of the assay. The bar indicates the median. **C-D.** Neutralizing titers are presented as the log 10 of the dilution inhibiting 50% of target infection.

In striking contrast with infected patients, the humoral response of the vaccinated group against RBD was heterogeneous, since most patients (17/29, 59%) failed to generate detectable levels of anti-RBD IgG after 2 doses of vaccine (**Figure 3B**). This defect was even more patent when considering the functional assay, in which only 21% of vaccinated patients had neutralizing humoral response against the pseudovirus (6/29; **Figure 3C**).

These findings led us to hypothesize that the lack of protection against COVID-19 in some vaccinated transplant recipients may be due to insufficient generation of neutralizing anti-RBD antibodies. To test this theory, we retrieved the 14 available serum samples collected after the 2 doses of mRNA-1273 but prior COVID-19 diagnosis for the vaccinated patients of the first cohort. In line with our theory, almost all these patients (12/14, 86%; **Figure 3E**) had no anti-RBD IgG after the “standard” scheme of vaccination and, consequently, their serum was unable to block the entry of pseudo-virus in human cells in neutralization assay (**Figure 3E**).

### Generation of neutralizing antibodies after vaccination requires spike-specific Tfh cells

The generation of IgGs against a protein antigen (such as spike) depends upon a prototypical T-cell-dependent humoral response. Following the binding of the antigen to their surface immunoglobulin (which delivers the first signal of activation), spike-specific B cell clones indeed require a second signal of activation that comes from cognate interactions with CD4+ T follicular helper (Tfh) cells (*32–34*).

Because our group (*9, 32*) and others (*35*) have shown that maintenance immunosuppression heterogeneously impacts Tfh population in transplant recipients, we speculated that a Tfh defect could explain the lack of generation of neutralizing antibodies observed in some vaccinated patients.

To test this theory, the 29 vaccinated transplant recipients were distributed into the group “responder” (n=12/29, 41%) or “non-responder” (n=17/29, 59%) according to whether or not they had detectable level of anti-RBD IgG in their circulation after 2 doses of mRNA-1273 vaccine. Clinical and biological characteristics of these two groups are presented in **Table 3** and were similar, except for age that was lower in the patients of the “responder” group.

**Table 3.** Characteristics of vaccinated patients.

| n (%) or median [IQR] | Non-responders<br>N = 17 | Responders<br>N = 12 | p* |
| --- | --- | --- | --- |
| **Age (y)** | 66.0 [50.2; 71.9] | 51.7 [41.0; 62.6] | **0.038** |
| **Male** | 11 (65) | 7 (58) | 0.728 |
| **Time since transplantation (y)** | 8.1 [1.6 ; 16.6] | 5.7 [2.7; 14.8] | 0.879 |
| **Immunosuppressive drugs** |  |  | 0.232 |
| CNI | 16 (94) | 12 (100) | 0.999 |
| MMF/MPA | 15 (88) | 7 (58) | 0.092 |
| Steroids | 11 (65) | 8 (67) | 0.913 |
| imTOR | 1 (6) | 2 (17) | 0.939 |
| Belatacept | 1 (6) | 0 (0) | 0.393 |
| **Biological data** |  |  |  |
| Lymphocytes (G/L) | 1.24 [1.0; 1.8] | 1.29 [1.0; 2.0] | 0.922 |
| Monocytes (G/L) | 0.55 [0.40; 0.74] | 0.56 [0.47; 0.86] | 0.639 |
| CRP (mg/L) | 4.0 [4.0; 8.4] | 4.0 [4.0; 5.2] | 0.884 |
| Albumin (g/L) | 43 [42; 46] | 44 [43; 46] | 0.502 |
| Creatinine (μmol/L) | 122 [90; 183] | 135 [111; 222] | 0.584 |

\*: qualitative variables were compared using a Chi-square test, quantitative variables were compared using a Mann-Whitney test.
**Abbreviations:** IQR: interquartile range; y, years; CNI: calcineurin inhibitor; MMF/MPA: mycophenolate mofetil/mycophenolic acid; imTOR, inhibitor of the mechanistic target of rapamycin.

Although Tfh cells act in B cell area within secondary lymphoid organs, recent studies have demonstrated that human blood CXCR5^+^CD4^+^ T cells are counterparts of Tfh and contain specific subsets that differentially support antibody secretion and can be identified on the basis of their profile of chemokine receptor expression (*36*). In line with these previous works, the 3 subsets of Tfh: Tfh1 (CXCR3^+^CCR6^-^), Tfh2 (CXCR3^-^ CCR6^-^), Tfh17 (CCR6^+^), could be identified and enumerated by flow cytometry in the circulation of vaccinated patients (**Figure 4A**). No difference was observed regarding the global count of CD4+ T cells, Tfh, or any of the Tfh subsets between responders and non-responders (**Figure 4B**). However, in line with our theory, spike-specific Tfh 1 and Tfh17 clones, which are the most efficient to drive antibody generation (*9, 36*), were found in higher quantity in the circulation of responders than non-responders (Tfh1: 3033±4570 vs 664±859 cells/mL, p=0.024; and Tfh17: 1010±1269 vs 306±385 cells/mL, p=0.020 in responders vs non responders; **Figure 4C**).

**Figure 4:**
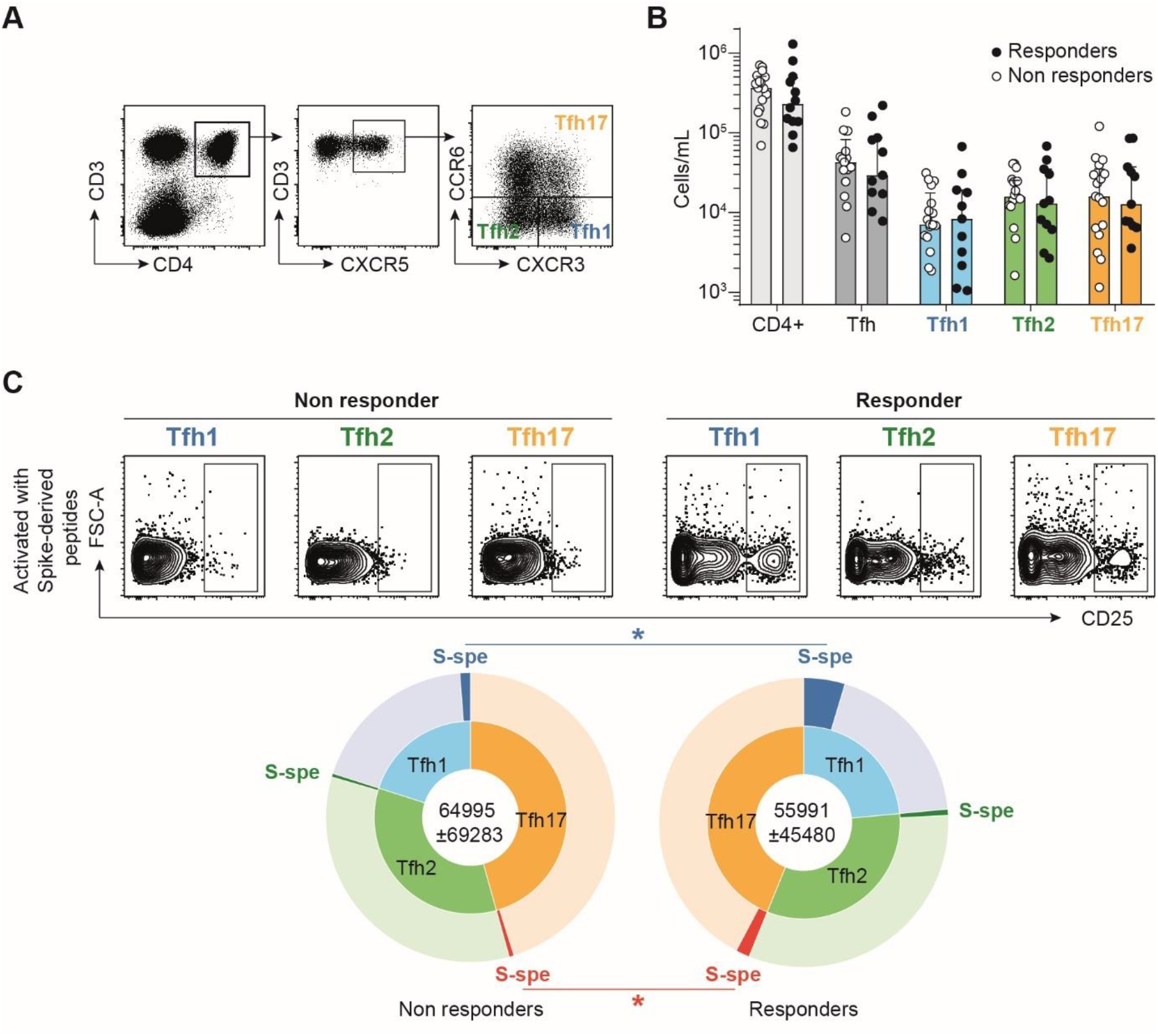
Generation of neutralizing antibodies after vaccination requires spike-specific Tfh cells. Follicular helper T cells (Tfh) were enumerated in the circulation of responders (black circle) and non-responders (open circle) to 2 doses of SARS-CoV-2 mRNA vaccine. **A.** Representative flow cytometry profiles with the gating strategy used to identify the 3 subsets of follicular helper T cells (Tfh): Tfh1 (blue), Tfh2 (green), and Tfh17 (orange). **B.** The count of various circulating CD4+ T cells subsets is plotted for each patient. Median are shown. Multiple t-tests, p>0.05. **C.** Spike-specific cells were enumerated among each Tfh subsets for each vaccinated patient. Sunburst charts were used to compare non-responders (left) and responders (right). Multiple t-tests; *, p<0.05.

### High mycophenolate mofetil dose and reduced vaccine immunogenicity impede spike-specific Tfh generation

Basic immunology works have demonstrated that Tfh number is a major limiting checkpoint that regulates the dynamic of germinal center reaction (*37, 38*). The reduced count in spike-specific Tfh therefore provides a likely explanation to the lack of generation of anti-RBD IgG in non-responders to vaccine. What could be the reason for this problem? Among the immunosuppressive drugs used in maintenance regimen, some block the activation of T cells (calcineurin-inhibitor) while other act by blocking the proliferation of adaptive immune effectors (mycophenolate mofetil). While responders and non-responders to vaccine were similarly exposed to calcineurin-inhibitors, non-responders received significantly more mycophenolate mofetil (500mg/day, IQR [0,1000] vs 1000mg/day, IQR [500; 1000] in responders vs non responders, p=0.027; **Figure 5A**).

**Figure 5:**
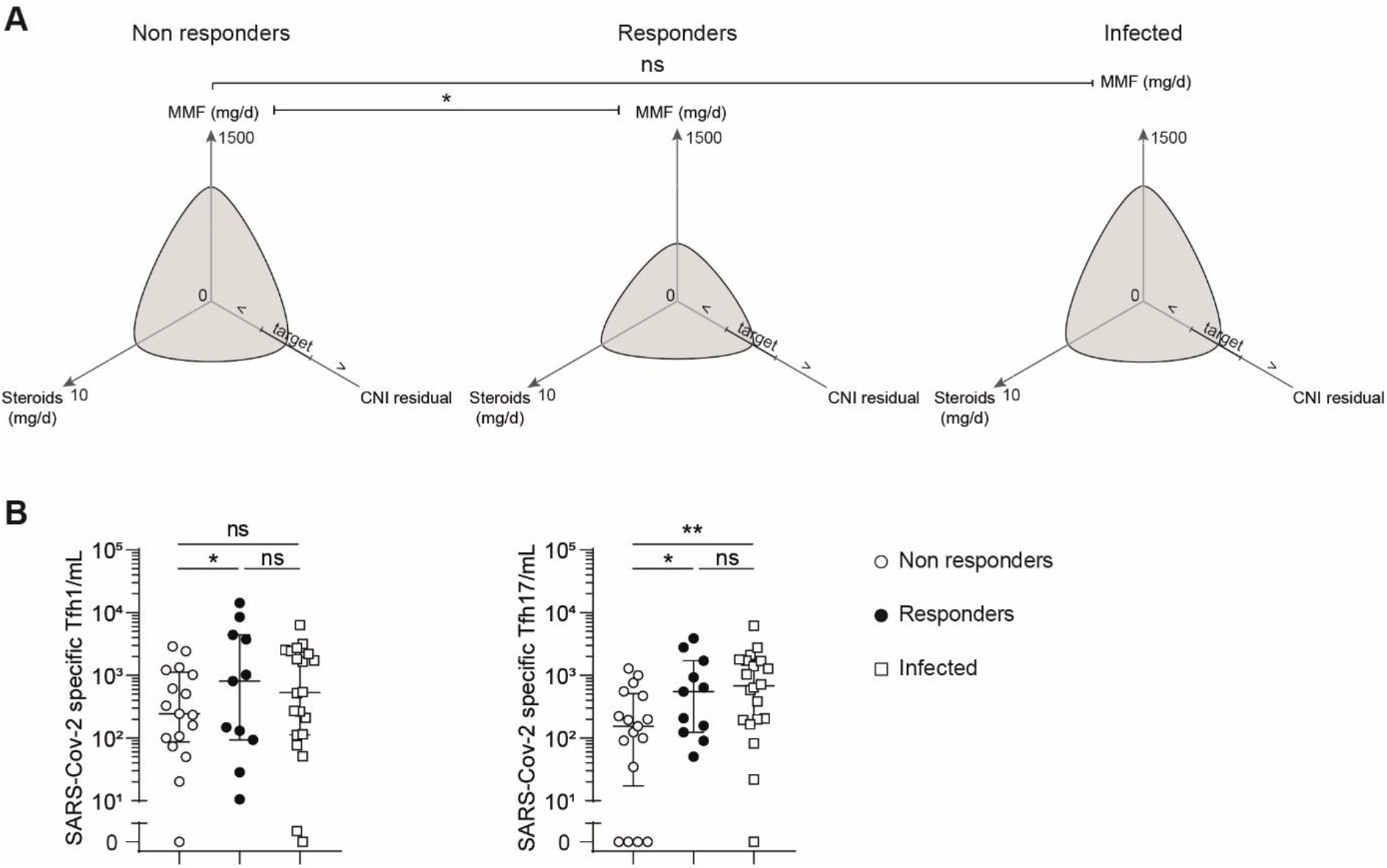
High mycophenolate mofetil dose impedes SARS-CoV-2-specific Tfh generation. **A.** Polar plots were used to compare maintenance immunosuppression regimen of non-responders (left panel) and responders (middle panel) to 2 doses of SARS-CoV-2 mRNA vaccine, and patients infected with SARS-CoV-2 (right panel). Median are plotted. Mann-Whitney test; *, p≤0.05. **B.** The count of SARS-CoV-2-specific Tfh1 and Tfh17 cells enumerated in the circulation of each non-responders (open circle) and responders (black circle) to 2 doses of SARS-CoV-2 mRNA vaccine, and patients infected with SARS-CoV-2 (open square) are plotted. The bar indicates the median. Multiple t-tests; ns, p>0.05; *, p≤0.05, **, p<0.01.

This result suggests that the anti-proliferative effect of high dose mycophenolate mofetil is the cause of the lack of response after 2 doses of mRNA-1273 vaccine observed in some transplant recipients. However, despite the fact that infected patients received the same high dose of mycophenolate mofetil at the time of infection as non-responders to vaccine (**Figure 5A**), they generated the same number of virus-specific Tfh1 and Tfh17 (**Figure 5B**), and consequently similar levels of neutralizing anti-RBD IgG (**Figure 3B & 3C**), as responders to vaccine.

### Increasing mRNA vaccine immunogenicity with a 3^rd^ dose improves anti-RBD IgG response of transplant patients

Our last observation led us hypothesizing that the detrimental impact of high dose mycophenolate mofetil could be overcome by a stronger/more immunogenic stimulation than the vaccine, such as the one provided to the patients by infection with live virus. In line with this hypothesis, vaccinated patients without anti-RBD IgG after 2 doses of mRNA-1273 did generate anti-RBD IgG after infection (**Figure 6A**).

**Figure 6:**
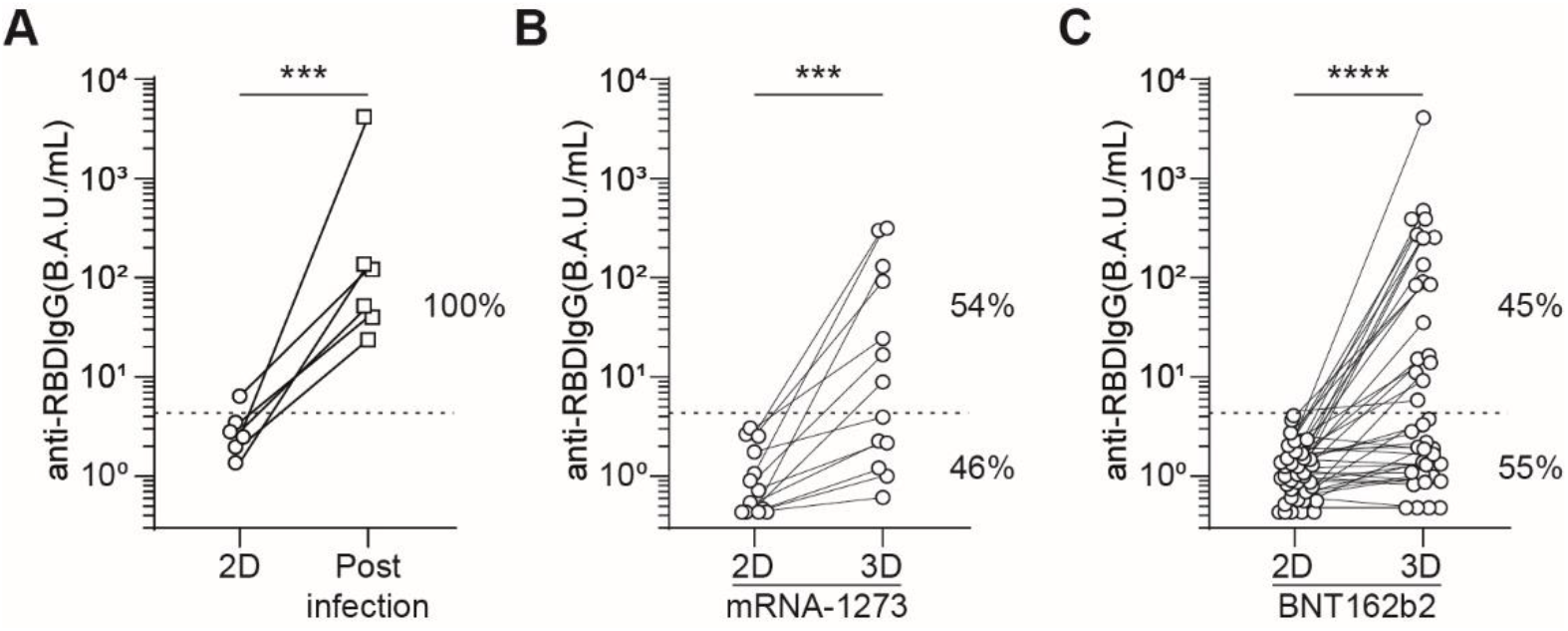
Increasing immunogenicity improves anti-RBD IgG response. **A.** Comparison of anti-RBD IgG titers measured after 2 doses (2D) of mRNA vaccine and after infection by SARS-CoV-2 in the same non-responders to vaccine transplant recipients. **B-C.** Comparison of anti-RBD IgG titers measured after the second (2D) and third (3D) dose of mRNA vaccine in the same patients, non-responders to standard scheme of vaccination. **B.** Discovery cohort (mRNA-1273 vaccine). **C.** External validation cohort (BNT162b2 vaccine). Wilcoxon test; ***, p<0.001, ****, p<0.0001.

Based on these results, we went on testing the impact of an additional dose of vaccine on the anti-RBD IgG response of 13 of the 17 transplant patients that were non-responders to the “standard” 2 doses scheme of vaccination with mRNA-1273. In accordance with our hypothesis, we observed that 7/13 (54%) did develop anti-RBD IgG after the 3^rd^ dose of vaccine (**Figure 6B**).

Aiming at validating this finding in an external cohort, a 3^rd^ dose of BNT162b2 mRNA vaccine was injected to a cohort of 36 renal transplant recipients from Lyon University hospital without detectable anti-RBD IgG after 2 injections. In accordance with our previous results, about half (15/36, 42%; **Figure 6C**) of these non-responders developed anti-RBD IgG after the 3^rd^ dose of vaccine.

## Discussion

While antibody levels are emerging as correlates of protection against COVID-19 in healthy subjects (*39*), there is still an urgent need to understand the relative contribution of humoral and T cell immunity in conferring protection to immunosuppressed populations (*17*), in particular transplant patients, who are both at high risk of death due to COVID-19 (*3–7*) and poor responders to mRNA vaccines (*11–14*).

Taking advantage of an original observation: i.e., that a previous infection by SARS-CoV-2 but not the standard “2 doses” scheme of vaccination, provided protection against COVID-19 to transplant recipients, we designed this translational study to compare the adaptive immune responses of these 2 groups of patients. This approach identified the generation of anti-RBD-IgG as a critical component of the adaptive immune response associated with protection against COVID-19 in transplant recipients. Generation of switched (IgG) antibodies against a protein antigen depends upon the germinal center, which is known to be controlled by Tfh cells, a subset of CD4+ T cells that act as the major limiting checkpoint of this reaction (*37, 38*). In line with these basic immunology works, non-responders to SARS-CoV-2 mRNA vaccine (i.e. transplant patients without anti-RBD-IgG after 2 doses of vaccine) were indeed characterized by reduced numbers of spike-specific Tfh cells. This was likely due to higher exposure to mycophenolate mofetil, an immunosuppressive drug that act by blocking the proliferation of activated lymphocytes (*40, 41*). This theory is supported by other independent studies, which have also observed an association between exposition to mycophenolate mofetil and lower antibody responses (*35, 42, 43*), including to SARS-CoV-2 vaccines (*11, 14*). Based on these findings, it is tempting to speculate that a reduction (or suspension) of the maintenance dose of mycophenolate mofetil prior vaccination might help obtaining better response rates. On the other hand, this non-antigen specific attitude might also favor the expansion of donor-specific Tfh, and therefore increase the risk of generation of *de novo* donor-specific antibodies (*9*), which is the first cause of late allograft loss (*44*) through accelerated chronic vascular rejection (*45, 46*).

Based on the observation that infected patients successfully generated anti-RBD IgG despite high dose of mycophenolate mofetil, similar to that of non-responders, we hypothesized that increasing immunogenicity of vaccine could allow improving patients’ protection without reducing maintenance immunosuppression. In line with this hypothesis, administration of a third dose of mRNA vaccine indeed resulted in the generation of anti-RBD IgG in about half of non-responders to standard scheme of vaccination. This result, was further validated in a larger independent prospective cohort with the other approved SARS-CoV-2 mRNA vaccines and has just been reported by independent groups (*47, 48*).

In addition to increasing the number of injections, another possibility to increase vaccine immunogenicity is to increase the amount of antigen provided in each injection. This strategy has been successfully tested in transplant recipients with protein-based vaccines against influenza (*49, 50*). However, it is unclear whether (and to which extend) increasing the doses of mRNA would result in an increased production of the pathogen-derived protein antigen by patient’s cells with these new vaccines platforms. To the best of our knowledge, there is currently no data in the literature addressing this issue.

At last, adaptation of vaccination scheme has limits and there will probably remain a fraction of transplanted patients that cannot generate an efficient antibody response whatever the vaccination scheme. In the latter, protection against COVID-19 might depend on infusion of mAb (or cocktails of mAb). This primary prevention strategy has indeed been successfully tested with bamlanivimab on another vulnerable population: 966 residents and staff in assisted living facilities (*51*). In this study mAb infusion halved the incidence of COVID-19 in the prevention population compared with placebo and the 5 deaths attributed to COVID-19 all occurred in the placebo group. Further studies, evaluating this strategy of passive immunization in organ transplant recipients are currently being discussed and their results are impatiently expected by transplant physicians.

## Patients and methods

### Study populations

#### Incidence of SARS-CoV-2 infection in kidney transplant recipients

The incidence of SARS-CoV-2 infections was monitored since the beginning of the pandemic, in the entire cohort of kidney transplant recipients at the University Hospital of Strasbourg (France), and compared between patients with a previous history of COVID-19 and those who received the 2 doses “standard” scheme of vaccination with mRNA-1273. The protection conferred by mRNA vaccine is operant as early as 12 days after the first injection in the general population (18). Transplant recipients were therefore considered “vaccinated” if they had received the second dose of vaccine. Kaplan-Meier’s method was used to compare COVID-19 incidence in the 2 populations. Data were censored at either date of death or June 16, 2021.

#### COVATRHUS cohort (COvid-19 VAccine in Transplant Recipients, Hopitaux Universitaires de Strasbourg)

To analyze the immune mechanisms involved in protection against COVID-19, 29 patients, naive for SARS-CoV-2 infection, were prospectively recruited from the cohort of kidney transplant recipients of the University Hospital of Strasbourg (France). According to the recommendations of the French health authority, they received a standard (2 doses) vaccination with mRNA-1273 (Moderna) COVID-19 vaccine. A third vaccine injection of mRNA-1273 COVID-19 vaccine was offered to all patients who did not develop IgG against the receptor binding domain of spike viral protein (anti-RBD IgG) after the second dose.

Vaccinated patients were compared to 21 patients retrospectively recruited among adult kidney transplant recipients of the University Hospital of Strasbourg, (France) who were diagnosed with COVID-19 between November 1, 2020 and January 31, 2021. The diagnosis of COVID-19 was based on positive testing of nasopharyngeal swabs by reverse transcription-polymerase chain reaction (RT-PCR). The study protocol complied with the tenets of the Helsinki Declaration and was approved by the Institutional Review Board (approval number: 18/21 03, Comité de Protection des Personnes Ouest IV Nantes) and registered on clinicaltrial.gov as NCT04757883.

Clinical, demographic, and laboratory data were collected at the time of the first vaccine injection or at the time of the COVID-19 diagnosis. Severity of COVID-19 was graded as asymptomatic, mild, moderate, severe, critical or death following the WHO recommendations (*19*).

The immune response after vaccination or infection was assessed at day 14 after the second dose of vaccine or one month after symptoms onset, respectively.

#### External validation cohort for the 3^rd^ dose of mRNA vaccine

This external validation cohort consisted of non-responders to 2 doses of BNT162b2 vaccine (Pfizer-BioNtech) from the cohort of kidney transplant recipients of Lyon University Hospital (France). The study protocol was approved by the local Institutional Review Board (approval number: 2020-A02918-31).

### Assessment of cellular immune responses directed against SARS-CoV-2

Peripheral Blood Mononuclear Cells (PBMC) were collected and isolated by centrifugation on a Ficoll density gradient. The cells were then frozen in fetal calf serum supplemented with 10% dimethylsulfoxide (DMSO, Sigma).

The SARS-CoV-2 specific CD8+ T cells and CD4+ T cells were identified as previously described (*9, 20*). Briefly, after thawing, cells were concentrated at 10^7 cells/mL in RPMI complete medium and left to rest overnight at 37°C and 5% CO2 in a 96-well round-bottom plate, 10^6 cells/well. The next day, the RPMI medium was changed, and the cells were cultured for 24 hours in the presence of peptide pools derived from the viral proteins spike, nucleocapsid and membrane (PepMix^TM^, JPT Peptides Technologies GmbH, Berlin, Germany). The pools contained overlapping peptides covering the entire sequence of the viral protein antigen. The final concentration of the peptides was 1µg/mL. Cells cultured with DMSO (Sigma) alone (1/250) were used as negative controls. Cells were then rinsed and incubated at room temperature with the relevant fluorescent antibodies for 30 minutes: CD3 (UHCT1), CD8 (SK1), CXCR3 (1C6), CXCR5 (RF8B2), CCR6 (11A9), CD25 (2A3), from BD Biosciences; CD4 (SK3), CD69 (FN50), CD137 (4B4-1), from Biolegend; and a Fixable Viability Dye (eBiosciences). Cells were fixed with 2% methanol-free formaldehyde. Sample acquisitions were made on a BD LSR Fortessa 4L flow cytometer (BD Biosciences).

### Assessment of humoral immune responses directed against SARS-CoV-2

#### Anti-Spike RBD IgG

IgG directed against the Receptor Binding Domain (anti-RBD IgG) of the spike glycoprotein of the SARS-CoV-2 were detected by a chemiluminescence technique, using the Maglumi® SARS-CoV-2 S-RBD IgG test (Snibe Diagnostic, Shenzen, China) on a Maglumi 2000® analyser (Snibe Diagnostic), according to the manufacturer’s instructions. This test displays clinical sensitivity and specificity of 100% and 99.6%, respectively.

Following WHO recommendation (*21*), the titers are expressed as binding antibody units/mL (BAU/mL); correction factor for Maglumi®: 4.33.

### Anti-nucleocapsid IgG antibodies

The Abbott anti-N IgG assay is an automated chemiluminescence microparticle immunoassay (CMIA) conducted and interpreted according to manufacturer guidelines. A sample-to-calibrator relative light unit index of ≥1.4 is considered positive, an index of ≥0.49 to <1.40 is considered borderline, and an index of <0.49 is considered negative. This CMIA displays clinical sensitivity and specificity of 96.5% and 99.2%, respectively (*22*).

### Neutralization assays

The assay was performed as follows: 3×10^4 293T-ACE2 (provided by O. Schwartz Laboratory, Institut Pasteur) were plated in 96-well plates. Sera were sequentially diluted and incubated with D614G Spike-pseudotyped lentiviral particles (provided by Rossolillo Laboratory, IGBMC) for 1 hour at +37°C. The mix were added to cells. After 72h, the intracellular luciferase signal was measured with Bright Glo luciferase assay system by a luminescence Counter MicroBetaTriLux 1450LSC (Perkin Elmer). The percentage of neutralization was calculated as: 100 x (1-(mean(luciferase signal in sample duplicate))/(mean(luciferase signal in virus alone))). The results are reported as the log_10_ of the dilutions that inhibit 50% of the infection of the targets [log_10_(IC50)].

### Statistical analysis

All the analyses were carried out using GraphPad Prism v8.0 (San Diego, California USA). Qualitative variables were expressed as percentages and compared with the chi-squared test, or Fisher’s exact test when the conditions of application of chi-square were not respected. Quantitative variables were expressed as mean ± SD and compared using multiple t-tests or as median ± IQR and compared using Mann-Whitney test for variables with non-normal distribution. Paired data were compared using Wilcoxon test. Incidence data analyzed by the Kaplan-Meier’s method were compared using a Log-rank test. A non-linear regression was performed to study the correlation of continuous quantitative variables.

## Data Availability

not applicable

## Acknowledgements

The authors are indebted to Audrey Kochman and the members of the GRoupe de REcherche Clinique (GREC: Céline Dagot, Farah Pauwels, Fatiha M’Raiagh and Daniel Sperandio) for excellent technical assistance during the collection of the samples. OT is thankful to Lise Siard, Claudine Lecuelle, and Philippe Favre from Eurofins Biomnis for their help during the conduction of the study.

## Funding

XC is supported by a funding from the Société Francophone de Transplantation.

ME is supported by the Hospices civils de Lyon and currently holds a “poste accueil” position funded by INSERM.

OT is supported by the Etablissement Français du Sang and the Fondation pour la Recherche Médicale (PME20180639518).

## Author contribution

Experiment’s conception and design: XC, ME, OT

Acquisition of clinical samples: XC, ME, IB, FH, FB, GGV, MD, PP, AK, NC, CL, LM, EM, SC, OT

Realization of experiments: XC, ME, IB, FG, PR, ES, FP, AO

Data analysis: XC, ME, IB, AO, SFK, SC, OT

Writing – original draft: XC, OT

Writing – review and editing: XC, FH, FB, GGV, MD, PP, AK, NC, CL, FG, LM, PR, EM, TD, SFK, SC, OT

## Competing interests

Authors declare that they have no competing interests.

## Notes

### Competing Interest Statement

The authors have declared no competing interest.

### Clinical Trial

NCT04757883

### Funding Statement

XC is supported by a funding from the Societe Francophone de Transplantation. 
ME is supported by the Hospices civils de Lyon and currently holds a poste accueil position funded by INSERM. 
OT is supported by the Etablissement Francais du Sang and the Fondation pour la Recherche Medicale (PME20180639518).

### Author Declarations

The study protocol complied with the tenets of the Helsinki Declaration and was approved by the Institutional Review Board (approval number: 18/21 03, Comite de Protection de Personnes Ouest IV Nantes) and registered on clinicaltrial.gov as NCT04757883.

## References

1. I. N. Zhu, D. Zhang, W. Wang, X. Li, B. Yang, J. Song, X. Zhao, B. Huang, W. Shi, R. Lu, P. Niu, F. Zhan, X. Ma, D. Wang, W. Xu, G. Wu, G. F. Gao, W. Tan, China Novel Coronavirus Investigating and Research Team, A Novel Coronavirus from Patients with Pneumonia in China, 2019. N Engl J Med. 382, 727–733 (2020).

2. COVID-19 situation update worldwide, as of week 26, updated 8 July 2021. European Centre for Disease Prevention and Control, (available at https://www.ecdc.europa.eu/en/geographical-distribution-2019-ncov-cases).

3. E. J. Williamson, A. J. Walker, K. Bhaskaran, S. Bacon, C. Bates, C. E. Morton, H. J. Curtis, A. Mehrkar, D. Evans, P. Inglesby, J. Cockburn, H. I. McDonald, B. MacKenna, L. Tomlinson, I. J. Douglas, C. T. Rentsch, R. Mathur, A. Y. S. Wong, R. Grieve, D. Harrison, H. Forbes, A. Schultze, R. Croker, J. Parry, F. Hester, S. Harper, R. Perera, S. J. W. Evans, L. Smeeth, B. Goldacre, Factors associated with COVID-19-related death using OpenSAFELY. Nature. 584, 430–436 (2020).

4. S. Caillard, D. Anglicheau, M. Matignon, A. Durrbach, C. Greze, L. Frimat, O. Thaunat, T. Legris, V. Moal, P. F. Westeel, N. Kamar, P. Gatault, R. Snanoudj, A. Sicard, D. Bertrand, C. Colosio, L. Couzi, J. M. Chemouny, C. Masset, G. Blancho, J. Bamoulid, A. Duveau, N. Bouvier, N. Chavarot, P. Grimbert, B. Moulin, Y. Le Meur, M. Hazzan, French SOT COVID Registry, An initial report from the French SOT COVID Registry suggests high mortality due to COVID-19 in recipients of kidney transplants. Kidney Int. 98, 1549–1558 (2020).

5. K. J. Jager, A. Kramer, N. C. Chesnaye, C. Couchoud, J. E. Sánchez-Álvarez, L. Garneata, F. Collart, M. H. Hemmelder, P. Ambühl, J. Kerschbaum, C. Legeai, M. D. Del Pino Y Pino, G. Mircescu, L. Mazzoleni, T. Hoekstra, R. Winzeler, G. Mayer, V. S. Stel, C. Wanner, C. Zoccali, Z. A. Massy, Results from the ERA-EDTA Registry indicate a high mortality due to COVID-19 in dialysis patients and kidney transplant recipients across Europe. Kidney Int. 98, 1540–1548 (2020).

6. O. Thaunat, C. Legeai, D. Anglicheau, L. Couzi, G. Blancho, M. Hazzan, M. Pastural, E. Savoye, F. Bayer, E. Morelon, Y. Le Meur, O. Bastien, S. Caillard, French nationwide Registry of Solid Organ Transplant Recipients with COVID-19, IMPact of the COVID-19 epidemic on the moRTAlity of kidney transplant recipients and candidates in a French Nationwide registry sTudy (IMPORTANT). Kidney Int. 98, 1568–1577 (2020).

7. S. Caillard, N. Chavarot, H. Francois, M. Matignon, C. Greze, N. Kamar, P. Gatault, O. Thaunat, T. Legris, L. Frimat, P. F. Westeel, V. Goutaudier, M. Jdidou, R. Snanoudj, C. Colosio, A. Sicard, D. Bertrand, C. Mousson, J. Bamoulid, C. Masset, A. Thierry, L. Couzi, J. M. Chemouny, A. Duveau, V. Moal, G. Blancho, P. Grimbert, A. Durrbach, B. Moulin, D. Anglicheau, Y. Ruch, C. Kaeuffer, I. Benotmane, M. Solis, Y. LeMeur, M. Hazzan, F. Danion, French SOT COVID Registry, Is COVID-19 infection more severe in kidney transplant recipients? Am J Transplant. 21, 1295–1303 (2021).

8. Vaccins Covid-19 : quelle stratégie de priorisation à l’initiation de la campagne ? Haute Autorité de Santé, (available at https://www.has-sante.fr/jcms/p_3221237/fr/vaccins-covid-19-quelle-strategie-de-priorisation-a-l-initiation-de-la-campagne).

9. A. S. Dahdal, C. Saison, M. Valette, E. Bachy, N. Pallet, B. Lina, A. Koenig, G. Monneret, T. Defrance, E. Morelon, O. Thaunat, Residual Activatability of Circulating Tfh17 Predicts Humoral Response to Thymodependent Antigens in Patients on Therapeutic Immunosuppression. Front Immunol. 9, 3178 (2018).

10. A. Duchini, J. A. Goss, S. Karpen, P. J. Pockros, Vaccinations for adult solid-organ transplant recipients: current recommendations and protocols. Clin Microbiol Rev. 16, 357–364 (2003).

11. B. J. Boyarsky, W. A. Werbel, R. K. Avery, A. A. R. Tobian, A. B. Massie, D. L. Segev, J. M. Garonzik-Wang, Antibody Response to 2-Dose SARS-CoV-2 mRNA Vaccine Series in Solid Organ Transplant Recipients. JAMA. 325, 2204–2206 (2021).

12. B. H. Rincon-Arevalo, M. Choi, A.-L. Stefanski, F. Halleck, U. Weber, F. Szelinski, B. Jahrsdörfer, H. Schrezenmeier, C. Ludwig, A. Sattler, K. Kotsch, A. Potekhin, Y. Chen, G. R. Burmester, K.-U. Eckardt, G. M. Guerra, P. Durek, F. Heinrich, M. Ferreira-Gomes, A. Radbruch, K. Budde, A. C. Lino, M.-F. Mashreghi, E. Schrezenmeier, T. Dörner, Impaired humoral immunity to SARS-CoV-2 BNT162b2 vaccine in kidney transplant recipients and dialysis patients. Sci Immunol. 6, eabj1031 (2021).

13. A. Sattler, E. Schrezenmeier, U. A. Weber, A. Potekhin, F. Bachmann, H. Straub-Hohenbleicher, K. Budde, E. Storz, V. Proß, Y. Bergmann, L. M. Thole, C. Tizian, O. Hölsken, A. Diefenbach, H. Schrezenmeier, B. Jahrsdörfer, T. Zemojtel, K. Jechow, C. Conrad, S. Lukassen, D. Stauch, N. Lachmann, M. Choi, F. Halleck, K. Kotsch, Impaired humoral and cellular immunity after SARS-CoV2 BNT162b2 (Tozinameran) prime-boost vaccination in kidney transplant recipients. J Clin Invest, 150175 (2021).

14. I. Benotmane, G. Gautier-Vargas, N. Cognard, J. Olagne, F. Heibel, L. Braun-Parvez, J. Martzloff, P. Perrin, B. Moulin, S. Fafi-Kremer, S. Caillard, Low immunization rates among kidney transplant recipients who received 2 doses of the mRNA-1273 SARS-CoV-2 vaccine. Kidney Int. 99, 1498–1500 (2021).

15. C. S. Caillard, N. Chavarot, D. Bertrand, N. Kamar, O. Thaunat, V. Moal, C. Masset, M. Hazzan, P. Gatault, A. Sicard, J. M. Chemouny, J. P. Rerolle, C. Colosio, H. Francois, J. Bamoulid, N. Bouvier, A. Duveau, D. Anglicheau, G. Blancho, French Society of Transplantation, Occurrence of severe COVID-19 in vaccinated transplant patients. Kidney Int, S0085-2538(21)00509–3 (2021).

16. D. N. M. Ali, N. Alnazari, S. A. Mehta, B. Boyarsky, R. K. Avery, D. L. Segev, R. A. Montgomery, Z. A. Stewart, Development of COVID-19 Infection in Transplant Recipients After SARS-CoV-2 Vaccination. Transplantation (2021), doi:10.1097/TP.0000000000003836.

17. E. P. S. Heeger, C. P. Larsen, D. L. Segev, Implications of defective immune responses in SARS-CoV-2 vaccinated organ transplant recipients. Science Immunology. 6 (2021), doi:10.1126/sciimmunol.abj6513.

18. F. L. R. Baden, H. M. El Sahly, B. Essink, K. Kotloff, S. Frey, R. Novak, D. Diemert, S. A. Spector, N. Rouphael, C. B. Creech, J. McGettigan, S. Khetan, N. Segall, J. Solis, A. Brosz, C. Fierro, H. Schwartz, K. Neuzil, L. Corey, P. Gilbert, H. Janes, D. Follmann, M. Marovich, J. Mascola, L. Polakowski, J. Ledgerwood, B. S. Graham, H. Bennett, R. Pajon, C. Knightly, B. Leav, W. Deng, H. Zhou, S. Han, M. Ivarsson, J. Miller, T. Zaks, COVE Study Group, Efficacy and Safety of the mRNA-1273 SARS-CoV-2 Vaccine. N Engl J Med. 384, 403–416 (2021).

19. COVID-19 Clinical management: living guidance, (available at https://www.who.int/publications-detail-redirect/WHO-2019-nCoV-clinical-2021-1).

20. A. Grifoni, D. Weiskopf, S. I. Ramirez, J. Mateus, J. M. Dan, C. R. Moderbacher, S. A. Rawlings, A. Sutherland, L. Premkumar, R. S. Jadi, D. Marrama, A. M. de Silva, A. Frazier, A. F. Carlin, J. A. Greenbaum, B. Peters, F. Krammer, D. M. Smith, S. Crotty, A. Sette, Targets of T Cell Responses to SARS-CoV-2 Coronavirus in Humans with COVID-19 Disease and Unexposed Individuals. Cell. 181, 1489–1501.e15 (2020).

21. A. G. Mattiuzzo, E. M. Bentley, M. Hassall, S. Routley, V. Bernasconi, P. Kristiansen, H. Harvala, D. Roberts, G. Semple, L. C. Turtle, P. J. Openshaw, K. Baillie, C. Investigators, L. S. H. Nissen-Meyer, A. B. Brants, E. Atkinson, P. Rigsby, D. Padley, N. Almond, N. J. Rose, M. Page, Establishment of the WHO International Standard and Reference Panel for anti-SARS-CoV-2 antibody, 60.

22. B. H. Wang, D. Wiredja, L. Yang, P. L. Bulterys, C. Costales, K. Röltgen, J. Manalac, J. Yee, J. Zehnder, R. Z. Shi, S. D. Boyd, B. A. Pinsky, Case-Control Study of Individuals with Discrepant Nucleocapsid and Spike Protein SARS-CoV-2 IgG Results. Clinical Chemistry. 67, 977–986 (2021).

23. C. R. A. Seder, P. A. Darrah, M. Roederer, T-cell quality in memory and protection: implications for vaccine design. Nat Rev Immunol. 8, 247–258 (2008).

24. D. R. Channappanavar, C. Fett, J. Zhao, D. K. Meyerholz, S. Perlman, Virus-specific memory CD8 T cells provide substantial protection from lethal severe acute respiratory syndrome coronavirus infection. J Virol. 88, 11034–11044 (2014).

25. E. M. J. Bevan, Helping the CD8(+) T-cell response. Nat Rev Immunol. 4, 595–602 (2004).

26. F. X. Chen, R. Li, Z. Pan, C. Qian, Y. Yang, R. You, J. Zhao, P. Liu, L. Gao, Z. Li, Q. Huang, L. Xu, J. Tang, Q. Tian, W. Yao, L. Hu, X. Yan, X. Zhou, Y. Wu, K. Deng, Z. Zhang, Z. Qian, Y. Chen, L. Ye, Human monoclonal antibodies block the binding of SARS-CoV-2 spike protein to angiotensin converting enzyme 2 receptor. Cell Mol Immunol. 17, 647–649 (2020).

27. G. R. Shi, C. Shan, X. Duan, Z. Chen, P. Liu, J. Song, T. Song, X. Bi, C. Han, L. Wu, G. Gao, X. Hu, Y. Zhang, Z. Tong, W. Huang, W. J. Liu, G. Wu, B. Zhang, L. Wang, J. Qi, H. Feng, F.-S. Wang, Q. Wang, G. F. Gao, Z. Yuan, J. Yan, A human neutralizing antibody targets the receptor-binding site of SARS-CoV-2. Nature. 584, 120–124 (2020).

28. H. Y. Wu, F. Wang, C. Shen, W. Peng, D. Li, C. Zhao, Z. Li, S. Li, Y. Bi, Y. Yang, Y. Gong, H. Xiao, Z. Fan, S. Tan, G. Wu, W. Tan, X. Lu, C. Fan, Q. Wang, Y. Liu, C. Zhang, J. Qi, G. F. Gao, F. Gao, L. Liu, A noncompeting pair of human neutralizing antibodies block COVID-19 virus binding to its receptor ACE2. Science. 368, 1274–1278 (2020).

29. I. E. Seydoux, L. J. Homad, A. J. MacCamy, K. R. Parks, N. K. Hurlburt, M. F. Jennewein, N. R. Akins, A. B. Stuart, Y.-H. Wan, J. Feng, R. E. Whaley, S. Singh, M. Boeckh, K. W. Cohen, M. J. McElrath, J. A. Englund, H. Y. Chu, M. Pancera, A. T. McGuire, L. Stamatatos, Analysis of a SARS-CoV-2-Infected Individual Reveals Development of Potent Neutralizing Antibodies with Limited Somatic Mutation. Immunity. 53, 98–105.e5 (2020).

30. B. Ju, Q. Zhang, J. Ge, R. Wang, J. Sun, X. Ge, J. Yu, S. Shan, B. Zhou, S. Song, X. Tang, J. Yu, J. Lan, J. Yuan, H. Wang, J. Zhao, S. Zhang, Y. Wang, X. Shi, L. Liu, J. Zhao, X. Wang, Z. Zhang, L. Zhang, Human neutralizing antibodies elicited by SARS-CoV-2 infection. Nature. 584, 115–119 (2020).

31. J. M. Espi, X. Charmetant, T. Barba, L. Koppe, C. Pelletier, E. Kalbacher, E. Chalencon, V. Mathias, A. Ovize, E. Cart-Tanneur, C. Bouz, L. Pellegrina, E. Morelon, D. Fouque, L. Juillard, O. Thaunat, The ROMANOV study found impaired humoral and cellular immune responses to SARSCov-2 mRNA vaccine in virus unexposed patients receiving maintenance hemodialysis. Kidney International. 0 (2021), doi:10.1016/j.kint.2021.07.005.

32. C.-C. Chen, A. Koenig, C. Saison, S. Dahdal, G. Rigault, T. Barba, M. Taillardet, D. Chartoire, M. Ovize, E. Morelon, T. Defrance, O. Thaunat, CD4+ T Cell Help Is Mandatory for Naive and Memory Donor-Specific Antibody Responses: Impact of Therapeutic Immunosuppression. Front Immunol. 9, 275 (2018).

33. A. Lanzavecchia, Antigen-specific interaction between T and B cells. Nature. 314, 537–539 (1985).

34. S. Crotty, Follicular helper CD4 T cells (TFH). Annu Rev Immunol. 29, 621–663 (2011).

35. S. Heidt, D. L. Roelen, C. Eijsink, M. Eikmans, C. van Kooten, F. H. J. Claas, A. Mulder, Calcineurin inhibitors affect B cell antibody responses indirectly by interfering with T cell help. Clin Exp Immunol. 159, 199–207 (2010).

36. R. Morita, N. Schmitt, S.-E. Bentebibel, R. Ranganathan, L. Bourdery, G. Zurawski, E. Foucat, M. Dullaers, S. Oh, N. Sabzghabaei, E. M. Lavecchio, M. Punaro, V. Pascual, J. Banchereau, H. Ueno, Human blood CXCR5(+)CD4(+) T cells are counterparts of T follicular cells and contain specific subsets that differentially support antibody secretion. Immunity. 34, 108–121 (2011).

37. T. A. Schwickert, G. D. Victora, D. R. Fooksman, A. O. Kamphorst, M. R. Mugnier, D. Gitlin, M. L. Dustin, M. C. Nussenzweig, A dynamic T cell-limited checkpoint regulates affinity-dependent B cell entry into the germinal center. J. Exp. Med. 208, 1243–1252 (2011).

38. M. C. Woodruff, E. H. Kim, W. Luo, B. Pulendran, B Cell Competition for Restricted T Cell Help Suppresses Rare-Epitope Responses. Cell Rep. 25, 321–327.e3 (2018).

39. S. Feng, D. J. Phillips, T. White, H. Sayal, P. K. Aley, S. Bibi, C. Dold, M. Fuskova, S. C. Gilbert, I. Hirsch, H. E. Humphries, B. Jepson, E. J. Kelly, E. Plested, K. Shoemaker, K. M. Thomas, J. Vekemans, T. L. Villafana, T. Lambe, A. J. Pollard, M. Voysey, the O. C. V. T. Group, medRxiv, in press, doi:10.1101/2021.06.21.21258528.

40. O. Thaunat, A. Koenig, C. Leibler, P. Grimbert, Effect of Immunosuppressive Drugs on Humoral Allosensitization after Kidney Transplant. J. Am. Soc. Nephrol. 27, 1890–1900 (2016).

41. A. C. Allison, E. M. Eugui, Immunosuppressive and other effects of mycophenolic acid and an ester prodrug, mycophenolate mofetil. Immunol Rev. 136, 5–28 (1993).

42. K. G. Smith, N. M. Isbel, M. G. Catton, J. A. Leydon, G. J. Becker, R. G. Walker, Suppression of the humoral immune response by mycophenolate mofetil. Nephrol Dial Transplant. 13, 160–164 (1998).

43. G. H. Struijk, R. C. Minnee, S. D. Koch, A. H. Zwinderman, K. A. M. I. van Donselaar-van der Pant, M. M. Idu, I. J. M. ten Berge, F. J. Bemelman, Maintenance immunosuppressive therapy with everolimus preserves humoral immune responses. Kidney Int. 78, 934–940 (2010).

44. R. S. Gaston, J. M. Cecka, B. L. Kasiske, A. M. Fieberg, R. Leduc, F. C. Cosio, S. Gourishankar, J. Grande, P. Halloran, L. Hunsicker, R. Mannon, D. Rush, A. J. Matas, Evidence for Antibody-Mediated Injury as a Major Determinant of Late Kidney Allograft Failure. Transplantation. 90, 68–74 (2010).

45. E. Pouliquen, A. Koenig, C. C. Chen, A. Sicard, M. Rabeyrin, E. Morelon, V. Dubois, O. Thaunat, Recent advances in renal transplantation: antibody-mediated rejection takes center stage. F1000Prime Rep. 7, 51 (2015).

46. O. Thaunat, Humoral immunity in chronic allograft rejection: Puzzle pieces come together. Transplant Immunology. 26, 101–106 (2012).

47. N. Kamar, F. Abravanel, O. Marion, C. Couat, J. Izopet, A. Del Bello, Three Doses of an mRNA Covid-19 Vaccine in Solid-Organ Transplant Recipients. N Engl J Med (2021), doi:10.1056/NEJMc2108861.

48. W. A. Werbel, B. J. Boyarsky, M. T. Ou, A. B. Massie, A. A. R. Tobian, J. M. Garonzik-Wang, D. L. Segev, Safety and Immunogenicity of a Third Dose of SARS-CoV-2 Vaccine in Solid Organ Transplant Recipients: A Case Series. Ann Intern Med (2021), doi:10.7326/L21-0282.

49. Y. Natori, M. Shiotsuka, J. Slomovic, K. Hoschler, V. Ferreira, P. Ashton, C. Rotstein, L. Lilly, J. Schiff, L. Singer, A. Humar, D. Kumar, A Double-Blind, Randomized Trial of High-Dose vs Standard-Dose Influenza Vaccine in Adult Solid-Organ Transplant Recipients. Clinical Infectious Diseases. 66, 1698–1704 (2018).

50. M. Mombelli, N. Rettby, M. Perreau, M. Pascual, G. Pantaleo, O. Manuel, Immunogenicity and safety of double versus standard dose of the seasonal influenza vaccine in solid-organ transplant recipients: A randomized controlled trial. Vaccine. 36, 6163–6169 (2018).

51. M. S. Cohen, A. Nirula, M. J. Mulligan, R. M. Novak, M. Marovich, C. Yen, A. Stemer, S. M. Mayer, D. Wohl, B. Brengle, B. T. Montague, I. Frank, R. J. McCulloh, C. J. Fichtenbaum, B. Lipson, N. Gabra, J. A. Ramirez, C. Thai, W. Chege, M. M. Gomez Lorenzo, N. Sista, J. Farrior, M. E. Clement, E. R. Brown, K. L. Custer, J. Van Naarden, A. C. Adams, A. E. Schade, M. C. Dabora, J. Knorr, K. L. Price, J. Sabo, J. L. Tuttle, P. Klekotka, L. Shen, D. M. Skovronsky, BLAZE-2 Investigators, Effect of Bamlanivimab vs Placebo on Incidence of COVID-19 Among Residents and Staff of Skilled Nursing and Assisted Living Facilities: A Randomized Clinical Trial. JAMA. 326, 46–55 (2021).

